# Homebrew reagents for low cost RT-LAMP

**DOI:** 10.1101/2021.05.08.21256891

**Authors:** Tamara Matute, Isaac Nuñez, Maira Rivera, Javiera Reyes, Paula Blázquez-Sánchez, Aníbal Arce, Alexander J. Brown, Chiara Gandini, Jennifer Molloy, César A. Ramirez-Sarmiento, Fernán Federici

## Abstract

RT-LAMP (reverse transcription - Loop-mediated isothermal amplification) has gained popularity for the detection of SARS-CoV-2. The high specificity, sensitivity, simple protocols and potential to deliver results without the use of expensive equipment has made it an attractive alternative to RT-PCR. However, the high cost per reaction, the centralized manufacturing of required reagents and their distribution under cold chain shipping limits RT-LAMP’s applicability in low-income settings. The preparation of assays using homebrew enzymes and buffers has emerged worldwide as a response to these limitations and potential shortages. Here, we describe the production of Moloney murine leukemia virus (M-MLV) Reverse Transcriptase and BstLF DNA polymerase for the local implementation of RT-LAMP reactions at low cost. These reagents compared favorably to commercial kits and optimum concentrations were defined in order to reduce time to threshold, increase ON/OFF range and minimize enzyme quantities per reaction. As a validation, we tested the performance of these reagents in the detection of SARS-CoV-2 from RNA extracted from clinical nasopharyngeal samples, obtaining high agreement between RT-LAMP and RT-PCR clinical results. The in-house preparation of these reactions results in an order of magnitude reduction in costs, and thus we provide protocols and DNA to enable the replication of these tests at other locations. These results contribute to the global effort of developing open and low cost diagnostics that enable technological autonomy and distributed capacities in viral surveillance.

## Introduction

Coronavirus disease 2019 (COVID-19) is a respiratory infection caused by the Severe Acute Respiratory Syndrome Coronavirus 2 (SARS-CoV-2)^1,2^. The detection of SARS-CoV-2 has relied primarily on quantitative reverse transcription-PCR (qRT-PCR) reaction^3,4^. Although qRT-PCR is the gold standard method for nucleic acid detection^5,6^, this technique requires sophisticated equipment and skilled personnel that has led to centralized strategies of COVID-19 diagnosis with high turnaround times. These limitations have encouraged the search for alternative techniques for the detection of SARS-CoV-2^7^. Among them, LAMP (Loop mediated isothermal amplification) has gained popularity due to its high specificity, sensitivity, simple protocols, more tolerance to inhibitors and potential to deliver results without the use of expensive instruments^8–10^. These advantages make it ideal for decentralized testing and surveillance of human pathogenic agents (e.g. Dengue^11^, MERS^11^, and HIV^12,13^).

LAMP relies on DNA polymerases with strand displacement capability, primarily the large fragment (exo-) pol I from *Geobacillus stearothermophilus* (formerly *Bacillus stearothermophilus*), called BstLF^14^. For RT-LAMP, the reaction is coupled with a reverse transcription step by adding a reverse transcriptase (e.g. HIV^15^ or RTx^16^) or utilizes a single enzyme with both activities (e.g. Bst3.0^17^).

Despite these advantages, the high cost of enzymes can pose significant challenges to deployment in low-resourced areas. Enzymes are usually manufactured in a few centralized facilities located in the global north and distributed worldwide under cold chain shipping, which cannot be always maintained. Although there are no indications of RT-LAMP reagent shortages, centralized production models are prone to the global supply chain disruptions that have affected access to other diagnostic resources during the COVID-19 pandemic^18,19^, further limiting the testing capacity in many countries of the global south. In response to these limitations, many groups have described the successful implementation of homemade RT-PCR and RT-LAMP protocols based on homebrewed enzymes.^15,20–28^

Here, we describe the in-house production of RT-LAMP reactions for SARS-CoV-2 from homebrewed Moloney murine leukemia virus (M-MLV) reverse transcriptase and Bst Large Fragment (BstLF) polymerase. We compare their performance to commercial reactions and tested their ability to detect SARS-CoV-2 from RNA extracted from clinical nasopharyngeal samples.

## Materials and methods

### Expression and purification of M-MLV and BstLF

The M-MLV gene was synthesized as a Gblock from IDT and cloned into at pET-19b vector with N-terminal 10x His-tag. BstLF DNA sequence from the OpenEnzyme collection was cloned into pET15b with a 6x His-tag at its N-terminus. The BstLF sequence in the Open Enzyme collection was obtained from *Geobacillus stearothermophilus* (GenBank U23149, UniProt P52026). The large fragment (LF) was selected between the amino acids 298-876 of the full length protein. The wild type BstLF sequence was then codon optimized for *E. coli* through the OPTIMIZER^29^ web server, using the *E. coli* codon usage from the HEG database and a guided random method. Sequences were verified by Sanger sequencing services from Eton Bio and full sequence files are provided here^30,31^. Both DNAs can be obtained from the Open Bioeconomy Lab and/or FreeGenes collection^32^. M-MLV and BstLF were expressed in *E. coli* BL21(DE3) and C41 respectively and purified following two open access protocols described in detail on the protocols.io Reagent Collaboration Network (ReClone) collection^33,34^. Briefly, protein production was induced with 0.5 mM IPTG for 16 hours at 18 ºC at 200 rpm after plasmid-containing *E. coli* cells grown in ampicillin-supplemented LB media reached OD_600_ = 0.8. Centrifugation-harvested cells were suspended in lysis buffer and disrupted first enzymatically using lysozyme and then with sonication on ice, using gentle cycles. Since BstLF is a thermostable protein, we added a thermolabile protein denaturation step by incubating the lysate at 65 ºC for 25 min. Lysates were then centrifuged at high speed to pellet cell debris and the supernatants were recovered and purified on Ni-NTA pre-packaged columns (HisTrap column, GE Healthcare). To remove imidazole, BstLF was buffer exchanged using Amicon Ultra-15 concentrators (Merck Millipore), whereas a second purification step using a HiTrap Heparin column (GE Healthcare) was employed for M-MLV. Protein concentration was determined by Bradford assay, using BSA as standard.

### RT-LAMP reaction setup

Buffers and reactions were prepared according to these online protocols (https://www.protocols.io/edit/low-cost-lamp-and-rt-lamp-bsejnbcn) deposited in the ReClone collection in protocols.io^35^. All reactions were performed on a StepOnePlus™ real time thermocycler (ThermoFisher) using the SYBR green filter. Reactions with commercial enzymes Bst2.0 WarmStart (NEB)^36^ and RTx (NEB)^36,37^ were performed according to the instructions from manufacturers. We used the primer set N2^38^ purchased from IDT (https://www.idtdna.com) with standard desalting.

A fragment of Nucleocapsid (N) gene RNA was used as positive control, which was produced by *in vitro* transcription from the IDT control for SARS-CoV-2 (catalogue 10006625). A PCR amplicon was first obtained using the following primers: NT7_Fw: CGA AAT TAA TAC GAC TCA CTA TAG GGG CAA CGC GAT GAC GAT GGA TAG; T7_Nter_Rv: ACT GAT CAA AAA ACC CCT CAA GAC CCG TTT AGA GGC CCC AAG GGG TTA TGC TAG TTA GGC CTG AGT TGA GTC AG, which added a T7 promoter and terminator to the amplicon. *In vitro* transcription was next performed from this linear DNA using the Hi-Scribe kit from NEB (catalogue E2040S) and treated with DNAse I (M0303S) for 15 minutes at 37°C before purification with Qiagen RNeasy kit, and serially diluted to the concentrations used.

The calculation of optimum enzyme concentrations was performed by maximizing an objective function which uses normalized functions for the amount of enzymes and the interpolation functions of normalized threshold time (nTt) and amplification step size (nSs). A higher weighting was assigned to nTt and nSs to compute the optimal enzyme values. Further procedures and computational routines are detailed in our github repository (https://github.com/LabTecLibres/RT-LAMP).

### Clinical samples

RNA samples were obtained by RNA extraction from nasopharyngeal clinical samples previously collected from anonymous patients that attended the outpatient service of Red Salud UC-CHRISTUS (Santiago, Chile). RNA was extracted by automated machines MagNA Pure (Roche) or ExiPrep™ (Bioneer Corp). Ct values of RT-PCR reactions were obtained with different primer sets for different samples, including CDC N1, N2, Orf1 and E sets. Samples with Ct values <= 40 were reported as positive SARS-CoV-2 cases.

### Data and materials availability

All raw data is provided in a Zenodo repository (https://zenodo.org/record/4540817#.YCnTO1NKj8o). DNAs can be obtained upon request and also from the FreeGenes collection^32^.

## Results and discussion

### Expression and purification of M-MLV and BstLF

The M-MLV gene was commercially synthesised from a design that contains point mutations D200N, L603W, T330P, L139P and E607K shown to increase thermostability and processivity^39^. The large fragment (LF) was synthesized from a sequence design that was codon optimized for *E. coli* from amino acids 298 to 876 of the wild type BstLF sequence from *Geobacillus stearothermophilus* (GenBank U23149, UniProt P52026).

BstLF and M-MLV were produced from IPTG inducible plasmids pET15b BstLF and pET-19b_MMLV-RT bearing 6x and 10x His-tags at the N-terminus, respectively, and expressed in *E. coli*, according to the open access protocols Rivera et al., 2020 ^33^ and Rivera et al., 2020b ^34^ (modified from Brown, 2020 ^40^). Then, the purification was performed in one Ni-NTA chromatography step for BstLF and in two chromatography steps (Ni-NTA and Heparin) for M-MLV, resulting in highly purified proteins in 3-4 days. Typical protein purification yields from 1 L cell culture correspond to 4 mg of M-MLV and 10 mg of BstLF.

### RT-LAMP reactions with homebrewed enzymes

To test the performance of the homebrewed enzymes, we compared RT-LAMP reactions composed of different combinations of BstLF, M-MLV and commercial enzymes Bst2.0 (NEB) and RTx (NEB) in their suggested concentrations; 0.32 U/μL and 0.3 U/μL, respectively (Fig. 1A). We tested the amplification of 10.000 copies of a RNA fragment of Nucleocapsid (N) gene of SARS-CoV-2, using the N2 LAMP primer set^38^. We found that 10.7 ng/μL of BstLF (and 62,5 pg/μL of M-MLV) to exhibit similar speed (i.e. time to threshold) as the commercial enzymes (i.e. Bst2.0 + RTx) whereas 21.3 ng/μL and 5.3 ng/μL to be faster and slower, respectively. We also tested different buffers. First, we tested two commercial buffers, Isothermal Amplification Buffer Pack (IA buffer)^41^ and ThermoPol® Reaction Buffer Pack (TPR Buffer)^42^, finding a better performance with IA buffer. Next, we compared homemade IA buffers with different concentrations of KCl: 50, 100 or 150 mM. Contrary to what was found in Aleeksenko et al., 2020^22^, we found that the IA buffer containing 50 mM KCl worked better than 100 and 150 mM (details in the online protocol). Mean time to threshold was 27.7 min for 50 mM of KCl, 43.5 min for 100 mM of KCl, and non-detectable for 150 mM of KCl. Therefore, homemade IA buffers containing 50 mM of KCl were used for the following assays.

**Fig 1.**
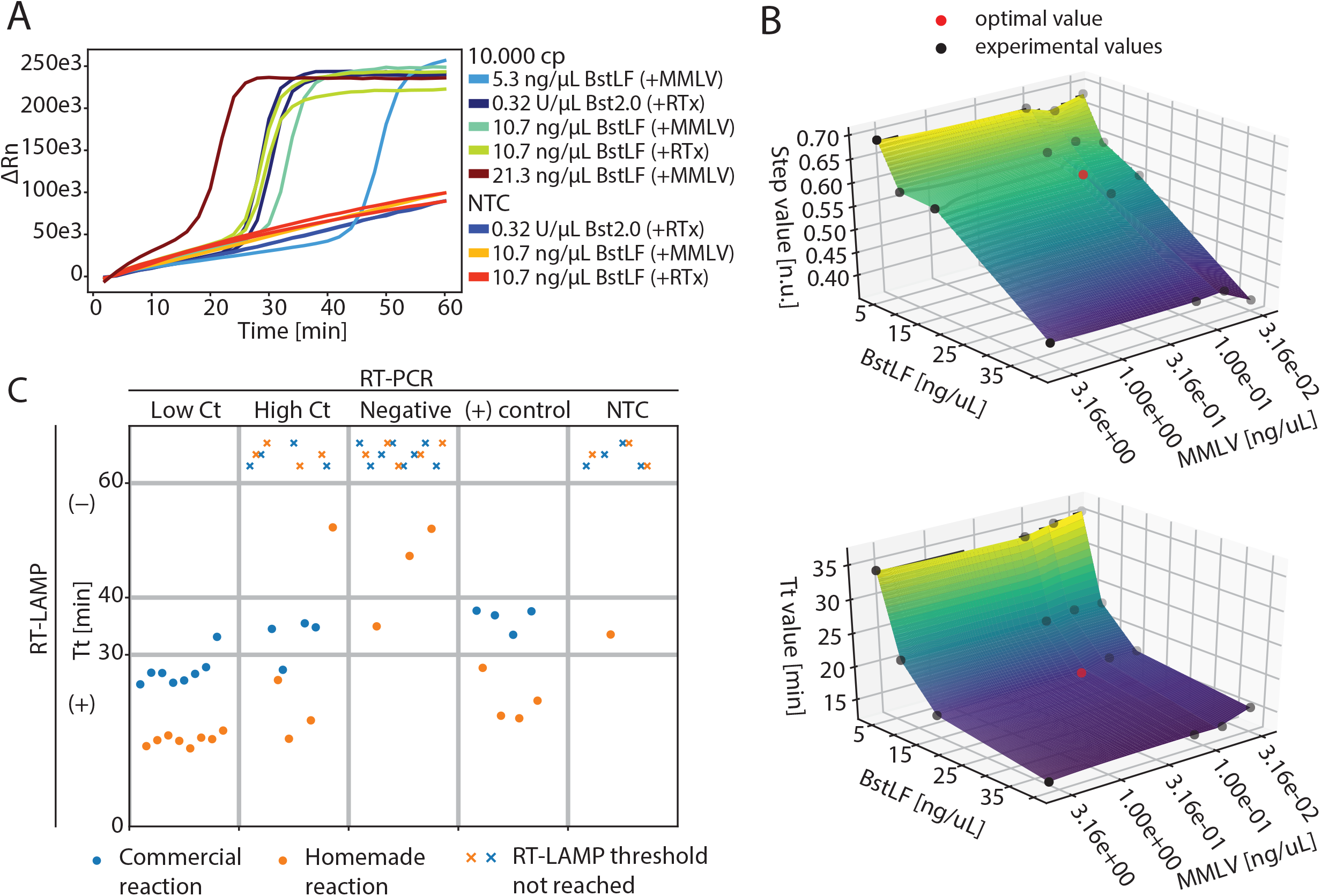
SARS-CoV-2 amplification with homemade RT-LAMP reactions. A) Comparison of RT-LAMP amplifications of 10000 copies of a RNA fragment from gene N using N2 primer set and the following enzyme combinations: BstLF + M-MLV, BstLF + RTx (NEB), and Bst2.0 (NEB) + RTx (NEB). M-MLV was employed at 62.5 pg/µL in all reactions while RTx was used at 0.3 U/µL as indicated by the manufacturer. NTC, no template control. B) Enzyme concentration optimization according to Tt, time to threshold (top), and step, ON/ OFF step (bottom). C) Detection of SARS-CoV-2 from RNA extracted from nasopharyngeal clinical samples. Positive controls correspond to 10.000 copies of a ssRNA fragment from the Nucleocapsid (N) gene. Threshold not reached label corresponds to RT-LAMP reactions that did not amplify its target during the whole time of testing. For the homemade reactions the mean Tt are 15 min for low Ct samples, 28 min for high Ct samples, 45 min for negative samples and 21 min for positive samples. For the commercial reactions the mean Tt are 27 min for low Ct samples, 33 min for high Ct samples, N/A for negative samples and 36 min for positive samples.

Next, we studied the effect of different concentrations of enzymes on time to threshold (Tt) and ON/OFF range (step) constrained by using as minimum amount of enzymes as possible in order to increase the number of reactions per purification (Fig. 1B). We found 16.52 ng/μL of BstLF and 0.127 ng/μL of M-MLV to be the optimum concentrations under these criteria. These concentrations were used for further assays and final tests with clinical samples.

### Homebrew reaction tested with clinical samples

To address the potential use of in-house prepared RT-LAMP reactions for the detection of SARS-CoV-2 from patient purified RNA, we tested 16 positive samples (8 of high and 8 of low Ct RT-PCR values with primer set N1 and N2) and 8 negative samples with NEB WarmStart commercial kit and the homemade BstLF/M-MLV RT-LAMP reactions. The samples corresponded to RNA extracted from nasopharyngeal clinical samples (Fig. 1C). Although both reactions, commercial and homemade, were able to detect high viral load samples (i.e. low Ct values), negative samples tested with commercial enzymes showed no amplification in any tube (0/8) whereas homemade enzymes exhibited one false positive (1/7). Samples with low viral load (i.e. high Ct values), on the other hand, were more likely to be classified as negatives with both systems (4 out of 8 in commercial reactions and 3 out of 8 in homemade reactions). A careful analysis of melting curves showed that the positive results presented the expected curve for the target, indicating non-spurious amplification of the specific sequence (data not shown but provided in the Zenodo repository link with raw data). The reactions from negative clinical samples (i.e. negative RT-PCR reaction) were classified as negative with the commercial RT-LAMP system whereas one tube was classified as positive in the homemade system. The melting curves showed that the positive result presented the expected curve of the target (i.e. not an amplification of spurious non-specific sequences), as it was the case for the two RT-LAMP negative results with Tt values between 40 and 60 min, indicating a possible cross-contamination from other tubes.

Positive control reactions were correctly classified as positives in both systems. Negative controls (NTC) were correctly classified with the commercial system whereas the homemade reaction misclassified one tube as positive. The melting curves showed that the positive result presented the expected curve of the target (i.e. not an amplification of spurious non-specific sequences), indicating a possible cross-contamination from another tube. Nevertheless, the time to threshold of false positives are clearly different from positive samples; indicating that a proper Tt selection (e.g. 30 min) should differentiate them. Although the analysis of melting curves suggests the absence of spurious unspecific amplifications, the homemade system provided faster times to threshold than the commercial reactions, which can be prone to deliver false negatives in reactions with high loads of non-target RNAs such as clinical samples^22^. Nevertheless, these results demonstrate that RT-LAMP reactions composed of in-house purified BstLF and M-MLV have comparable performance to commercial kits. The costs of consumables for the homemade reactions (USD 0.36 per reaction) is one order of magnitude cheaper than the costs of reactions with commercial enzymes (NEB WarmStart kit USD 3.86 per reaction and NEB RTx and Bst2.0 WarmStart enzymes bought separately USD 3.38 per reaction), enabling the development of more affordable alternatives for diagnostics. These results are in agreement with recent publications demonstrating the use of homebrewed enzymes for RT-LAMP detection of SARS-CoV-2 from patient samples^15,22,28^. These efforts have demonstrated the use of in-house produced BstLF and derivatives v7.16 and v5.9, along with a diverse set of homebrewed reverse transcriptases such as RTX (“xenopolymerase”)^28,43^, MashupRT (FeLV-RT-based)^22^ and HIV RT^15^. Here, we contribute with the use of in-house produced M-MLV for reverse transcription.

A variety of isothermal DNA amplification techniques have recently been developed and present themselves as highly versatile molecular biology tools to enable simple, in-the-field screening assays or point of care clinical diagnostics. Unlike traditional PCR, these reactions can be carried out at a constant temperature; eliminating the need for a thermocycler. Among these approaches is RT-LAMP, which has emerged as a promising and simple diagnostic tool for identifying SARS-CoV-2^9^. Advances on detection methods for LAMP reactions have described a series of alternatives with different degrees of equipment-dependency (or none), such as turbidity, gel electrophoresis, calcein, colorimetric indicators, pH-sensitive dyes, and DNA-intercalating fluorescent dyes (e.g. SYBR Green, SYTO dyes, EvaGreen)^44^. Our implemented protocol relies on the use of intercalating dye EvaGreen, as it has been shown to perform better than SYBR green^45,46^. A potential limitation of our proof of concept method is that the use of intercalating dyes can make target-specific real-time detection challenging^44^, and could pose as a barrier for use in resource limited areas. Future lower resource implementations of our method could substitute EvaGreen for 100-200 µM halochromic dyes, like Cresol Red, in pH 8.8 buffered reaction mix to cause detectable colorimetric change when amplification products have been produced^47^. Alternatively more advanced detection methods utilizing sequence specific methods based on the use of probes (e.g. LAMP-BEAC^23^, OSD^48^, DARQ^49^ and QUASR^50^) could also be utilized as end-point detection methods, and remain to be tested in future work. Such methods would be advantageous as they could be multiplexed for different targets in the same reaction. QUASR, for instance, has been shown to allow single step and close-tube multiplexing^50^.

The versions of both M-MLV and BstLF enzymes used in this paper are in the public domain in Chile but some of the M-MLV mutations are still subject to protection in other jurisdictions e.g. in the US (patent no. US7056716B2^51^) with anticipated expiration date of 15 March 2021. Future work will, therefore, explore the use of off-patent enzymes Open-MMLV or HIV RT^15^ from the *E. coli* expression toolkit from ReClone initiative^52^ in order to have freedom-to-operate in any country. The use of public domain enzymes would also permit commercial partnerships seeking to scale-up production locally. ReClone is a reagent collaboration network that addresses this challenge by providing access to open protocols and off-patent reagents for diagnostics under more permissive material transfer agreements (openMTA^53^), mitigating shortages as well as building preparedness in the long-term. Overall the Open RT-LAMP approach presented here demonstrates a viable method to replicate the detection sensitivities of traditional commercial RT-LAMP protocols at a fraction of the cost and with minimal resources. While we demonstrate the utility of the Open RT-LAMP protocol in the detection of SARS-CoV-2 RNA isolated from clinical samples, this approach could easily be applied to a broad range of screening assays to identify other infectious diseases or common pathogens at low cost. Our open RT-LAMP protocol aims to contribute to the global effort of making diagnostic capabilities more distributed, affordable and equitable.

## Data Availability

All data is available in the following repository: https://zenodo.org/record/4540817#.YCnTO1NKj8o

https://zenodo.org/record/4540817#.YCnTO1NKj8o

## Acknowledgments

This research was supported by ANID CONCYTEC covbio0012 and ANID – Millennium Science Initiative Program – ICN17_022. This work was further supported by the National Institutes of Health NIAID training grant (Training Program in Immunology; T32-AI07405) awarded to A. J. Brown. We thank the gLAMP consortium and the JOGL OpenCOVID community for the advice and for sharing relevant information for the establishment of LAMP reactions for SARS-CoV-2. We thank Carolina Miranda and Patricia Garcia from Red Salud UC-CHRISTUS for kindly providing PUC RNA samples from anonymous patients. We also thank Javier Gasulla (UBA, Argentina) for advice regarding the BstLF purification protocol. M. Rivera was funded by ANID FONDECYT 3190731 grant. P. Blázquez-Sánchez and J. Reyes were funded by ANID Doctoral Scholarship (ANID PFCHA 21191979 and 21191684, respectively).

## References

1. Naming the coronavirus disease (COVID-19) and the virus that causes it. https://www.who.int/emergencies/diseases/novel-coronavirus-2019/technical-guidance/naming-the-coronavirus-disease-(covid-2019)-and-the-virus-that-causes-it.

2. Zhu, N. et al. A Novel Coronavirus from Patients with Pneumonia in China, 2019. N. Engl. J. Med. 382, (2020).

3. Tang, Y.-W., Schmitz, J. E., Persing, D. H. & Stratton, C. W. Laboratory Diagnosis of COVID-19: Current Issues and Challenges. J. Clin. Microbiol. 58, (2020).

4. Website. https://www.fda.gov/media/134922/download.

5. Lu, X. et al. US CDC Real-Time Reverse Transcription PCR Panel for Detection of Severe Acute Respiratory Syndrome Coronavirus 2 - Volume 26, Number 8—August 2020 - Emerging Infectious Diseases journal - CDC. doi:10.3201/eid2608.201246.

6. Esbin, M. N. et al. Overcoming the bottleneck to widespread testing: A rapid review of nucleic acid testing approaches for COVID-19 detection. RNA rna.076232.120 (2020).

7. Guglielmi, G. The explosion of new coronavirus tests that could help to end the pandemic. Nature 583, 506–509 (2020).

8. Mini review: Recent progress in RT-LAMP enabled COVID-19 detection. Sensors and Actuators Reports 2, 100017 (2020).

9. Augustine, R. et al. Loop-Mediated Isothermal Amplification (LAMP): A Rapid, Sensitive, Specific, and Cost-Effective Point-of-Care Test for Coronaviruses in the Context of COVID-19 Pandemic. Biology 9, 182 (2020).

10. Notomi, T. et al. Loop-mediated isothermal amplification of DNA. Nucleic Acids Res. 28, E63 (2000).

11. Teoh, B.-T. et al. Detection of dengue viruses using reverse transcription-loop-mediated isothermal amplification. BMC Infect. Dis. 13, 1–9 (2013).

12. Rapid detection of human immunodeficiency virus type 1 group M by a reverse transcription-loop-mediated isothermal amplification assay. J. Virol. Methods 157, 195– 199 (2009).

13. Rapid detection of HIV-1 by reverse-transcription, loop-mediated isothermal amplification (RT-LAMP). J. Virol. Methods 151, 264–270 (2008).

14. Crystal structure of a thermostable Bacillus DNA polymerase l large fragment at 2.1 Å resolution. Structure 5, 95–108 (1997).

15. Kellner, M. J. et al. A rapid, highly sensitive and open-access SARS-CoV-2 detection assay for laboratory and home testing. Cold Spring Harbor Laboratory 2020.06.23.166397 (2020) doi:10.1101/2020.06.23.166397.

16. Zhang, Y. et al. Rapid Molecular Detection of SARS-CoV-2 (COVID-19) Virus RNA Using Colorimetric LAMP. medRxiv 2020.02.26.20028373 (2020).

17. New England Biolabs. Bst 3.0 DNA Polymerase. https://international.neb.com/products/m0374-bst-3-0-dna-polymerase#Product%20Information.

18. Nkengasong, J. Let Africa into the market for COVID-19 diagnostics. Nature 580, 565– 565 (2020).

19. Esfandiari, S. Wuhan, China, faces such a shortage of coronavirus test-kits that people say getting one is like ‘winning the lottery’. Business Insider France https://www.businessinsider.fr/us/wuhan-coronavirus-china-shortage-test-kits-lottery-2020-1 (2020).

20. Maranhao, A., Bhadra, S., Paik, I., Walker, D. & Ellington, A. D. An improved and readily available version of Bst DNA Polymerase for LAMP, and applications to COVID-19 diagnostics. medRxiv 2020.10.02.20203356 (2020).

21. Bhadra, S., Maranhao, A. C. & Ellington, A. D. One enzyme reverse transcription qPCR using Taq DNA polymerase. Cold Spring Harbor Laboratory 2020.05.27.120238 (2020) doi:10.1101/2020.05.27.120238.

22. Alekseenko, A. et al. Direct detection of SARS-CoV-2 using non-commercial RT-LAMP reagents on heat-inactivated samples. Sci. Rep. 11, 1–10 (2021).

23. Sherrill-Mix, S. et al. LAMP-BEAC: Detection of SARS-CoV-2 RNA Using RT-LAMP and Molecular Beacons. medRxiv 2020.08.13.20173757 (2020).

24. Bhadra, S., Riedel, T. E., Lakhotia, S., Tran, N. D. & Ellington, A. D. High-surety isothermal amplification and detection of SARS-CoV-2, including with crude enzymes. Cold Spring Harbor Laboratory 2020.04.13.039941 (2020) doi:10.1101/2020.04.13.039941.

25. Graham, T. G. W. et al. Inexpensive, versatile and open-source methods for SARS-CoV-2 detection. medRxiv 2020.09.16.20193466 (2020).

26. A blueprint for academic laboratories to produce SARS-CoV-2 quantitative RT-PCR test kits. J. Biol. Chem. 295, 15438–15453 (2020).

27. Vonesch, S. C. et al. McQ – An open-source multiplexed SARS-CoV-2 quantification platform. medRxiv 2020.12.02.20242628 (2020).

28. iSCAN: An RT-LAMP-coupled CRISPR-Cas12 module for rapid, sensitive detection of SARS-CoV-2. Virus Res. 288, 198129 (2020).

29. Puigbò, P., Guzmán, E., Romeu, A. & Garcia-Vallvé, S. OPTIMIZER: a web server for optimizing the codon usage of DNA sequences. Nucleic Acids Res. 35, W126–W131 (2007).

30. pET-19b_MMLV-RT· Benchling. https://benchling.com/s/seq-QR4HRtwI7IumNVoYKUeg.

31. pET15b_Bst-LF· Benchling. https://benchling.com/s/seq-5qNtNN9nE0HsQQLnX67N.

32. COVID-19 Diagnostic Toolkit Enzymes. https://stanford.freegenes.org/products/covid-19-diagnostic-toolkit-enzymes.

33. Rivera, M. Recombinant protein expression and purification of codon-optimized Bst-LF polymerase. (2020a) doi:10.17504/protocols.io.bksrkwd6.

34. Rivera, M. Recombinant expression and purification of codon-optimized M-MLV and Mashup. (2020b). https://dx.doi.org/10.17504/protocols.io.bsernbd6

35. Molloy, J. Reclone.org (The Reagent Collaboration Network). protocols.io https://www.protocols.io/groups/recloneorg-the-reagent-collaboration-network/recloneorg-the-reagent-collaboration-network/about (2020).

36. New England Biolabs. Bst 2.0 WarmStart DNA Polymerase. https://international.neb.com/products/m0538-bst-20-warmstart-dna-polymerase#Product%20Information.

37. New England Biolabs. WarmStart® RTx Reverse Transcriptase. https://international.neb.com/products/m0380-warmstart-rtx-reverse-transcriptase#Product%20Information.

38. Zhang, Y. et al. Enhancing colorimetric loop-mediated isothermal amplification speed and sensitivity with guanidine chloride. Biotechniques 69, (2020).

39. Baranauskas, A. et al. Generation and characterization of new highly thermostable and processive M-MuLV reverse transcriptase variants. Protein Eng. Des. Sel. 25, (2012).

40. Brown, A. Recombinant Protein Expression of MMLV-RT H (SkiBar H RT) V2 v2 (protocols.io.bijzkcp6). protocols.io doi:10.17504/protocols.io.bijzkcp6.

41. New England Biolabs. Isothermal Amplification Buffer Pack. https://international.neb.com/products/b0537-isothermal-amplification-buffer#Product%20Information.

42. New England Biolabs. ThermoPol® Reaction Buffer Pack. https://international.neb.com/products/b9004-thermopol-reaction-buffer#Product%20Information.

43. Ellefson, J. W. et al. Synthetic evolutionary origin of a proofreading reverse transcriptase. Science 352, 1590–1593 (2016).

44. Becherer, L. et al. Loop-mediated isothermal amplification (LAMP) – review and classification of methods for sequence-specific detection. Anal. Methods 12, 717–746 (2020).

45. Lee, S. H. et al. One-Pot Reverse Transcriptional Loop-Mediated Isothermal Amplification (RT-LAMP) for Detecting MERS-CoV. Front. Microbiol. 7, (2017).

46. Eischeid, A. C. SYTO dyes and EvaGreen outperform SYBR Green in real-time PCR. BMC Res. Notes 4, 1–5 (2011).

47. Tanner, N. A., Zhang, Y. & Evans, T. C., Jr. Visual detection of isothermal nucleic acid amplification using pH-sensitive dyes. Biotechniques 58, 59–68 (2015).

48. Jiang, Y. S. et al. Robust strand exchange reactions for the sequence-specific, real-time detection of nucleic acid amplicons. Anal. Chem. 87, (2015).

49. [No title]. https://www.future-science.com/doi/10.2144/0000113902?url_ver=Z39.88-2003&rfr_id=ori%3Arid%3Acrossref.org&rfr_dat=cr_pub++0pubmed.

50. [No title]. https://pubs.acs.org/doi/10.1021/acs.analchem.5b04054.

51. US 7056716 B2 - High Fidelity Reverse Transcriptases And Uses Thereof The Lens - Free & Open Patent and Scholarly Search. https://www.lens.org/lens.

52. Reagent Collaboration Network – a global collaboration for equitable access to biotechnology. https://reclone.org.

53. Kahl, L. et al. Opening options for material transfer. Nat. Biotechnol. 36, 923–927 (2018).

